# Factors influencing healthcare providers’ knowledge and skills retention following helping mothers survive and helping babies survive training in Tanzania: A mixed methods cross-sectional study

**DOI:** 10.1101/2024.11.19.24317563

**Authors:** Beatrice Erastus Mwilike, Martha Rimoy, Lucy Mabada, Nicodem Komba, Feddy Mwanga, Rashid Gosse, Joel Ambikile

**Author notes:** Corresponding author Beatrice Erastus Mwilike.

## Abstract

The Tanzanian Midwives Association (TAMA), in collaboration with development partners, implemented a project (*50,000 Happy Birthdays)* from 2018 to 2020 to improve providers’ knowledge and skills in saving lives at birth. The project was implemented under two training modules namely *Helping Mothers Survive* (HMS) and *Helping Babies Survive* (HBS). Through a mixed-methods cross-sectional design, knowledge retention was assessed by comparing follow-up and post-test knowledge scores among healthcare providers in the selected health facilities in Tanga, Geita, and Katavi regions. Four knowledge areas were evaluated using a written test and five skills areas were assessed using skills observation checklists for each. Quantitative data was analyzed using IBM SPSS version 25 by performing descriptive analysis, *t*-test and One-Way ANOVA with the level of significance determined at p< 0.05. Qualitative analysis was conducted through a thematic analysis approach and themes were generated to describe the factors influencing retention. 210 respondents participated in the study with more than half holding diploma (n=116; 55.2%) and working in urban area (n=123; 58.6%). There was a statistically significant drop in mean knowledge scores for controlling bleeding after birth (*t*=15.404, *p*<0.001), in helping babies breathe (*t*=8.580, *p*<0.001), and in essential care for small babies (*t*=19.620, *p*<0.001). Working in a rural area had a statistical significant higher drop in mean knowledge scores for managing pre-eclampsia (F=4.491, *p*=0.035) and for helping babies breathe (F=5.005, *p*=0.026). Education level also showed a significant difference in the mean knowledge score for managing pre-eclampsia (F=4.850, *p*=0.003).

There was poor knowledge retention following receiving training on HMS and HBS. The level of professional education and residential area significantly influenced knowledge retention. Frequent refresher training and other relevant training among healthcare workers may be helpful in knowledge retention regarding maternal and neonatal care, with much emphasis on providers working in rural areas and those with lower level of professional education.

## Introduction

Despite the global, regional, and local efforts to improve maternal and neonatal services, maternal and neonatal mortality have remained major public health issues for decades, particularly in low- and middle-income countries (LMICs). Globally, every two minutes, a woman dies from preventable causes related to pregnancy, which is statistically equivalent to approximately 800 maternal deaths per day [1]. Of these, almost 95% occur in LMICs [1, 2], with the limited skills of the birth attendants being one of the leading factors [2, 3]. Besides, approximately 6,500 newborns (within the first 28 days of birth) die every day. In 2022, 2.3 million newborns died in the first month of life, accounting for about 47 percent of all under-five deaths [4], mainly attributed to a lack of quality care at birth or skilled care and treatment immediately after birth and in the first days of life [5].

According to the 2022 Tanzania Demographic Health and Malaria Indicator Survey (TDHS - MIS), the maternal mortality ratio (MMR) was 104 maternal deaths per 100,000 live births, indicating a notable decline in MMR from 556 per 100,000 live births in 2015/2016 [6], and a step forward towards achieving the Standard Developmental Goal (SDG) 3.1 target of reducing MMR to less than 70 deaths per 100,000 live births by 2030 [7]. Unlikely, the neonatal mortality rate has been stagnant over the past two decades, decreasing at a slow pace from 40 deaths per 1,000 live births in 1999 to 25 deaths per 1,000 live births in 2015/2016 to 24 deaths per 1,000 live births in 2022 [6], indicating slow progress towards achieving the SDG 3.2.2 target of ending preventable newborn deaths by 2030 [7].

To reach the SDG 3.1 and 3.2.2 targets, deliberate analysis and corresponding interventions are needed across various settings, with extensive focus on those showing sluggishness in progress [6, 7]. Recently, the Tanzanian government has increased health infrastructure by building new health facilities and expanding the existing ones in an effort to improve accessibility to health services, including reproductive, maternal, newborn, and child health (RMNCH) services [8]. Subsequently, services such as caesarean sections, which were mainly provided at referral and tertiary hospitals, have been decentralized to primary healthcare settings, particularly district hospitals and health centers that meet the criteria. Despite these efforts, the skilled health personnel needed to provide quality services in these health facilities, as per the World Health Organization (WHO) recommendation, is one of the critical challenges [9, 10]. The WHO recommends high-quality care for all women during the antenatal, intranatal, and postnatal periods [11] and that all births be attended by skilled birth attendants (SBA) who are supplied with the necessary requirements, including in-service training [11, 12]. Yet, Tanzania faces an estimated half-shortage of skilled health workforce [13], and a notable shortage of SBA [14]. In fact, 66% of maternal deaths and 43% of newborn deaths can be prevented in births that occur with the assistance of SBAs who are supported and trained in basic life-saving competencies [15, 16], and one of the strategies to achieve the SDG 3.1 target by 2030 is to have at least 90% of births attended by SBA [7].

In response to this shortage and the need for high-quality services, in-service training has been implemented in multiple settings to strengthen the knowledge, skills, and performance of healthcare providers in maternal and newborn care units [17–20]. These trainings take into account various approaches and implementation frameworks, even though the content is quite similar [17, 21]. Of these, Helping Mothers Survive (HMS) and Helping Babies Survive (HBS) programs have been implemented as innovative training strategies that are cost-effective while meeting the demands for more basic and practical training, particularly in resource-limited settings [17, 22, 23]. It is mainly focused on the management of the leading contributors to maternal and neonatal deaths, including controlling bleeding after birth, active management of the third stage of labour, administration of magnesium sulphate, manual removal of the placenta, helping babies breathe, essential care for small babies, and feeding with a nasogastric tube [22, 23]. HMS and HBS training have shown varying effectiveness in the settings where they have been implemented. A review conducted in Tanzania reports a notable improvement in knowledge, skills, and performance among SBA and a reduction in early neonatal deaths and fresh stillbirths following in-job training on neonatal resuscitation [24]. Similarly, reviews from other regions have also highlighted post-training increases in knowledge, skills, and confidence [25, 26]. Even though, in terms of clinical practice, some studies have reported improvement [27–30], while others have reported no significant improvement [31]. It is further recommended that for better maternal and neonatal outcomes, multifaceted interventions with structured follow-up interventions are rather encouraged than single interventions or interventions with no follow-up [25, 26, 32].

Although HBS and HMS programs have shown significant impact in terms of improvement of knowledge and skills [28], post-training follow-up surveys show a decline in knowledge and skills [33, 34]. A training in rural Tanzania among SBA led to an improvement in knowledge, skills, and confidence immediately post-training [33], but a nine-month follow-up revealed a decline in knowledge and simulated skills [35]. Similarly, a cluster-randomised trial involving 61 facilities in Tanzania revealed a decline of skills to 4.0% at a 10-month follow-up of in-facility training on helping mothers survive bleeding after birth (HMS BAB) [34]. This entails the short-term retention of knowledge, skills, and competencies gained in training, thus necessitating the need to find out the factors that may be influencing post-training retention of knowledge and skills. Previous studies in other settings have highlighted factors such as frequency of training, years of clinical experience [36, 37], cadre variations [37, 38], and training modality [38]. However, they are not comprehensive enough to inform about knowledge and skill retention in our context.

Therefore, as we owe it to enhance the training outcomes in terms of long-term retention of knowledge and skills and subsequent improved clinical outcomes, our study aimed at exploring the factors influencing healthcare providers’ knowledge and skills retention following helping mothers survive and helping babies survive training in Tanzania, as they are not yet clear. In 2018 to 2020 the healthcare providers received training through a programme known as 50,000 Happy Birthdays (50KHB). The programme aimed to increase the professional capacity of midwives in three countries: Ethiopia, Rwanda and Tanzania. This was a post-training follow-up survey conducted among the healthcare providers who previously received HMS and HBS training mainly in four knowledge and five skills areas. We, therefore, aimed to assess the factors influencing competence retention following HMS and HBS in Tanzania.

## Materials and Methods

### Study setting

This study was conducted in six purposefully selected districts in three *50,000 Happy Birthday* project regions of mainland Tanzania namely Geita, Tanga and Katavi. The districts were Geita Town Council, Chato District Council, Tanga Town Council, Muheza District Council, Mpanda District Council and Mpanda Town Council. These districts were selected from regions where the 50KHB programme was conducted and impact evaluated. For each of the six districts, the team randomly selected three health facilities of different levels among those facilities which were part of the project. In each region, facilities were selected using the following criteria:

i. Facilities that have providers who are already trained on HMS and HBS modules.
ii. For the three regions selected, selected a mix of facilities that represent hospital, health center and dispensary.
iii. Facilities representing urban, and rural contexts from each region.

### Study design

Our study utilized a mixed-methods cross-sectional design to compare the current situation at selected sites with endline data from the *50,000 Happy Birthdays* project. Leveraging existing master trainers in each region for efficient and consistent data collection, the study employed standard *HMBS* evaluation documents – multiple-choice knowledge assessments and objective structured clinical evaluations (OSCEs) – as well as supplemental tools like facility-level interviews, clinical observations, and desk reviews to capture all relevant qualitative and quantitative information that may influence results.

We engaged a significant and representative sample of study participants to ensure that the findings can be applied to national best practices: we identified a diverse array of health facilities (district hospitals, health centres, dispensaries) and maternal/newborn service providers from each of the three project regions to evaluate. A subsequent, centralised analysis was conducted to understand common factors associated with competence retention among providers.

### Study Participants

This study involved two different populations: healthcare providers and tutors. Healthcare providers and tutors were enrolled from the following target populations:

**Target population 1** – Healthcare providers from selected health facilities who have been trained on HMS and HBS modules.

**Target population 2** – Supervisors, DRCHCOs and RRCHCOs, and educators who have been trained, and are proficient in the provision of maternal newborn and childcare in their contexts.

**Target population 3 –** Tutors who were trained on HMS and HBS modules in the selected Nursing and Midwifery Colleges.

### Inclusion and exclusion criteria

Healthcare providers (HCPs) and tutors were eligible for inclusion in the study if they were currently living in Geita, Katavi and Tanga regions, were classified as being one of the three target populations and had consented to participate in skills drills, focus groups or interviews. Participants were excluded if they were sick or on study leave and they plan to move from the region during the study period.

### Sampling Methods and Procedure Sampling for Quantitative Research

All the trained providers in the 28 purposively selected facilities and three training institutions were approached and enrolled in the study if they consented. Healthcare providers and tutors from selected health facilities who have been trained in HMS and HBS modules were recruited. Consecutive recruitment was employed until all the trained providers available were enrolled. Below in table 1 is the list of Health facilities and Training centers supported under 50KHB projects in Tanga, Geita, and Katavi regions. Overall, we were able to access a total of 210 participants from all the study sites **(Table 1).**

**Table 1.** Sample of the study sites.

|  | Tanga | Geita | Katavi |
| --- | --- | --- | --- |
| **District** | **Tanga Town Council** | **Geita Town Council** | **Mpanda Municipal Council** |
| **Education institution** | Tanga COHAS | Geita School of Nursing | n/a |
| **Hospital** | Bombo Regional Referral Hospital | Geita Regional Referral Hospital | Katavi Referral Regional Hospital |
| **Primary health facilities** | Ngamiani HC, Mikanjuni HC, Duga dispensary | Nyankumbu HC, Kasamwa HC, Bung'wankoko dispensary | Town council clinic, Kakese dispensary, Mwamkulu dispensary |
| **District** | **Muheza District Council** | **Chato District Council** | **Mpanda District council** |
| **Education institution** | St. Augustine Muheza IHAS | n/a | n/a |
| **Hospital** | Muheza DH | Chato DH | n/a |
| **Primary health facilities** | Mkuzi HC, Bulwa HC, Misozwe dispensary, Mkanyageni dispensary | Bwanga HC, Buzilamoyo dispensary, Butalama dispensary, Kibehe dispensary | Mwese HC, Mishamo HC, Kalema HC, Katuma dispensary, Sibwesa dispensary, Kasekeseke dispensary |

### Sampling for Qualitative research

The sampling of providers was purposive to meet the desirable characteristics. Providers were selected from diverse backgrounds to collate feedback from participants of different socio-demographic characteristics. Focus groups were arranged to include groups of similar characteristics. Participants included supervisors, DRCHCOs, RRCHCOs, and educators who have been trained and are proficient in the provision of maternal, newborn and childcare in their contexts.

The final sample sizes were based on theoretical saturation, which refers to the point at which no new concepts emerge from the review of data drawn from a sample that is diverse in pertinent characteristics and experiences. We achieved theoretical saturation after having three focus groups with target populations composed of 12 participants each. Also, we conducted a total of 16 in-depth interviews across all three regions.

### Data collection method and procedures

Data was collected from January to March 2023, approximately two years following the online survey for the 50,000 Happy Birthday project. The assessment of HCPs’ knowledge and skills utilized qualitative and quantitative approaches, including in-depth interviews, OSCE, and questionnaires. Participants were recruited from the three regions from 16 January until 31 March 2023.

**Qualitative data** from interviews and discussions were collected using an audio recorder. Data were then transcribed verbatim and translated from Kiswahili into English. For qualitative data collection, an interview guide will be used. Data were collected by a team of main researchers. The interviews lasted for about 30 to 60 minutes and were conducted in Kiswahili language.

**Quantitative data** were collected using the modified tools that were used for pre and posttest during the 50,000 Happy Birthday program intervention. The questionnaires were paper printed and participants were asked to fill in after providing signed informed consent. The researchers and research assistants were responsible for data collection.

Tools were prepared to assess the knowledge and skills retention in HMS and HBS among healthcare providers and tutors. The assessment focused on both knowledge and skills in managing two HMS modules (Bleeding After Birth Complete (BABC) and Pre-Eclampsia/Eclampsia (PEE)) and three HBS modules (Helping Babies Breathe (HBB2.0), Essential Care for Every Baby (ECEB) and Essential Care for Small Babies (ECSB)).

The knowledge test tools comprise of socio demographic information and multiple-choice questions for different topics covered during the training. The skills test tools comprise of socio demographic information, instructions for conducting the skills and observation checklists.

The knowledge test for HMS tools included a) Helping Mothers Survive: Bleeding after Birth Complete Knowledge Assessment b) Helping Mothers Survive: Knowledge Test Pre-Eclampsia & Eclampsia. The skills test for HMS tools included OSCE for a) Active Management of Third Stage of Labour (AMTSL) b) Shock Management, and c) Administering the loading dose of MgSO4. The knowledge test for HBS tools included a) Essential Care for Every Baby Knowledge Check b) Essential Care for Small Babies: Knowledge check. The skills test for HBS tools included a) Helping baby breath and b) Care for small babies.

### Data analysis

#### Quantitative data analysis

Quantitative data from questionnaires were transferred to an Excel database and later to Statistical Package for the Social Sciences (SPSS) version 25 for analysis. Descriptive analyses were conducted using frequency, central tendencies, and other relevant methods to establish follow up knowledge and skills retention. Similarly, these measures were used in determining the impact of the training program on improvement of the maternal and neonatal health outcomes. Data were compiled, cleaned, and analyzed for each specific objective as shown in the following Table 2.

**Table 2.**
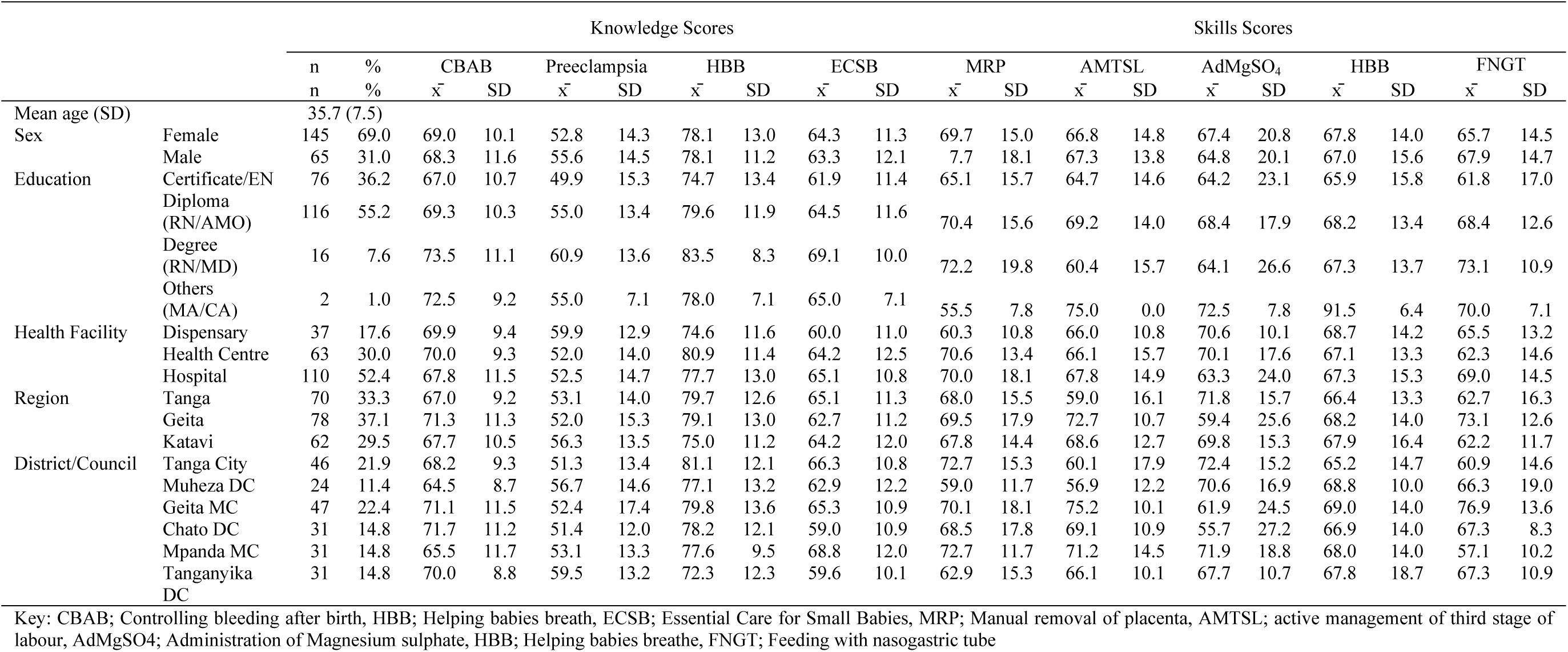
Demographic characteristics of participants and mean follow-up knowledge and skills scores for managing mothers and babies during and after child birth (n=210).

#### Qualitative data analysis

This study utilized deductive-inductive thematic analysis approach to identify, analyze, and interpret patterns within the qualitative data. The audio recording information was transcribed and transcripts in Swahili language were prepared. The researchers read through the translated scripts repetitively and compare them with the original audio record to check for accuracy. Thereafter, a list of meaningful information found was made and codes assigned. Then, the list was classified into themes and each theme was classified into sub-themes. Then, the data was translated and be presented in a text form. Validity in qualitative data based on triangulation, member checking, and collaboration with other investigators.

### Ethical approval

The ethical approval for this study was obtained from the Muhimbili University of Health and Allied Sciences Research and Ethics Committee with Ref.No.DA.282/298/01.C/. Permission to conduct the study was sought from the Ministry of Health (MoH), the President’s Office Regional Administration and Local Government (PORALG), and from regional and council/municipal authorities – with information being transmitted to facility authorities. Participation in this study was voluntary and participants were required to sign the written informed consent form. Time compensation was given to the participants after the interviews. The research team ensured that data remains confidential, and no identification will be published along with the reports of manuscripts. Interviews were conducted in isolated rooms to ensure confidentiality and data will be stored in locked cabinet in TAMA offices for a period of five years.

## Results

### Characteristics of the study participants

A total of 210 healthcare providers participated in the study, with the mean (SD) age of 35.7 (7.5), and most of them (n=145; 69.0%) were females. The majority (n=116; 55.2%) were diploma holders, followed by certificate holders (n=76; 36.2%), and just more than a half (n=110: 52.4%) were working in a regional or district referral hospital. The region with the highest number of participants was Geita (n=78; 37.1%) followed by Tanga (N=70: 33.3%), with Geita and Tanga municipal councils contributing to most participants (n=47; 22.4% and n=46: 21.9%, respectively). Regarding the residential area (not shown in table), more than half (n=123; 58.6%) of the participants were working in health facilities located in the urban area (**Table 2).**

### Participants’ knowledge retention scores

Four knowledge areas were assessed including, two for care of the mother i.e. how to control bleeding after childbirth and managing pre-eclampsia, and two for the care of the newborn i.e. helping babies to breathe and essential care for small babies. Participants’ mean follow-up knowledge (SD) score was highest for helping babies to breathe {78.1 (12.4)} and lowest for managing pre-eclampsia {53.6 (14.4)}. Follow-up knowledge scores were compared to post-test knowledge scores (previously assessed) and the change in mean (SD) scores was computed to determine knowledge retention. The results show that there was a drop in mean knowledge score in all areas assessed with the highest drop observed in managing pre-eclampsia {-21.0 (15.0)} and lowest in helping babies to breathe {-7.7 (13.0)}.

A paired *t*-test was performed to assess the differences between the follow-up mean knowledge scores and the post-test mean knowledge scores (**Table 3**) to determine whether the observed drop in knowledge scores were significant. For all knowledge areas assessed, there were statistically significant drop in knowledge scores. Specifically, there was a statistically significant drop in mean knowledge scores for controlling bleeding after birth (*t*=15.404, *p*<0.001), pre-eclampsia (*t*=20.276, *p*<0.001), helping babies breath (*t*=8.580, *p*<0.001), and essential care for small babies (*t*=19.620, *p*<0.001).

**Table 3.** Mean scores differences between follow-up and post-test knowledge scores.

| Area assessed | Post-test |  | Follow-up |  | Difference |  | Paired *t*-test |  |  |
| --- | --- | --- | --- | --- | --- | --- | --- | --- | --- |
|  | Mean | SD | Mean | SD | Mean | SD | 95%CI | *t* | p value |
| CBAB | 81.3 | 7.3 | 68.8 | 10.5 | -12.5 | 11.7 | 10.86703, 14.05678 | 15.404 | <0.001 |
| Pre-eclampsia | 74.6 | 8.1 | 53.6 | 14.4 | -21.0 | 15.0 | 18.93671, 23.01567 | 20.276 | <0.001 |
| HBB | 85.8 | 7.8 | 78.1 | 12.4 | -7.7 | 13.0 | 5.91977, 9.45166 | 8.580 | <0.001 |
| ECSB | 78.6 | 7.3 | 64.0 | 11.5 | -14.7 | 10.8 | 16.12989, 19.620 | 19.620 | <0.001 |

Key: CBAB; Controlling bleeding after birth, HBB; Helping babies breath, ECSB; Essential Care for Small Babies, CI; confidence interval

An independent *t*-test was performed to compare the mean knowledge drop scores between participants working in urban and rural areas. Participants working in rural area had a statistically significant higher drop in mean knowledge scores for managing pre-eclampsia (F=4.491, *p*=0.035) and for helping babies breath (F=5.005, *p*=0.026) **(Table 4).**

**Table 4.**
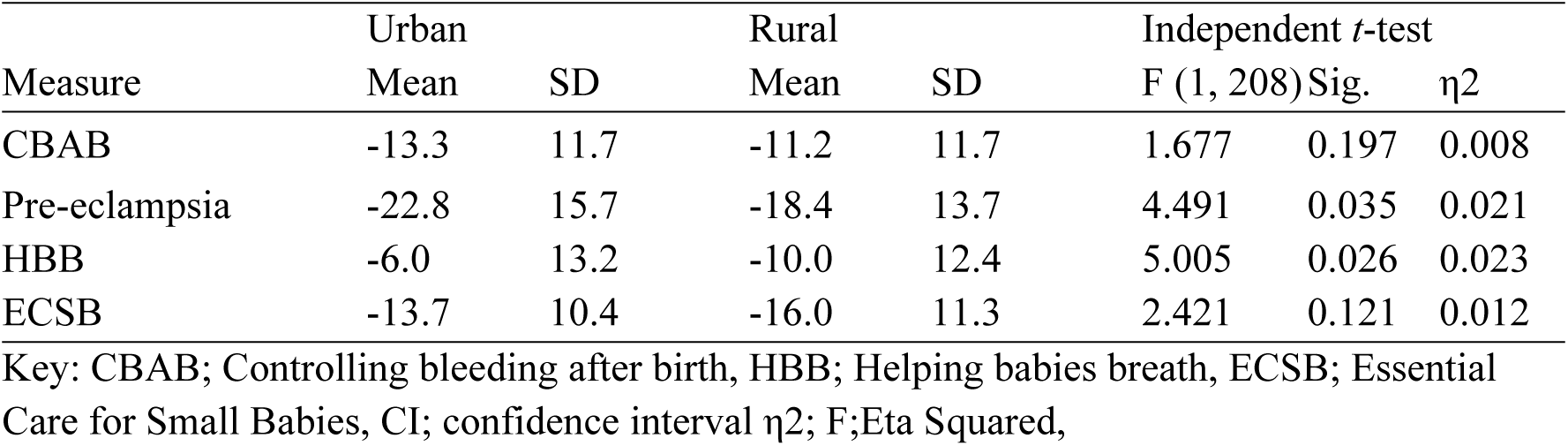
Univariate analysis of variance of knowledge according to residential area.

| Measure | Urban |  | Rural |  | Independent *t*-test |  |  |
| --- | --- | --- | --- | --- | --- | --- | --- |
| | Mean | SD | Mean | SD | F (1, 208) | Sig. | $\eta^2$ |
| CBAB | -13.3 | 11.7 | -11.2 | 11.7 | 1.677 | 0.197 | 0.008 |
| Pre-eclampsia | -22.8 | 15.7 | -18.4 | 13.7 | 4.491 | 0.035 | 0.021 |
| HBB | -6.0 | 13.2 | -10.0 | 12.4 | 5.005 | 0.026 | 0.023 |
| ECSB | -13.7 | 10.4 | -16.0 | 11.3 | 2.421 | 0.121 | 0.012 |

Key: CBAB; Controlling bleeding after birth, HBB; Helping babies breath, ECSB; Essential Care for Small Babies, CI; confidence interval $\eta^2$ ; F;Eta Squared,

Another statistical analysis that was performed for assessment of knowledge drop was the One-Way ANOVA to determine differences in mean knowledge scores among participants according to their levels of professional education i.e. holders of certificate, diploma, degree, and others (**Table 5**). There was a statistically significant difference only in the mean knowledge score for managing pre-eclampsia (F=4.850, *p*=0.003). When the Post Hoc Test was performed it was revealed that there were statistically significant mean knowledge differences in pre-eclampsia between certificate holders and diploma holders (*p*=0.006), and between certificate holders and degree holders (*p*=0.037). There were no significant differences in knowledge variance with other levels of professional education.

**Table 5.** One-Way ANOVA by level of professional education.

| Measure |  | Sum of squares | Mean square | F (3, 206) | p value |
| --- | --- | --- | --- | --- | --- |
| CBAB | Between Groups | 528.591 | 176.197 | 1.287 | 0.280 |
|  | Within Groups | 28197.604 | 136.882 |  |  |
|  | Total | 28726.195 |  |  |  |
| Pre-eclampsia | Between Groups | 3099.223 | 1033.074 | 4.850 | 0.003 |
|  | Within Groups | 43875.658 | 212.989 |  |  |
|  | Total | 46974.881 |  |  |  |
| HBB | Between Groups | 474.154 | 158.051 | 0.937 | 0.424 |
|  | Within Groups | 34745.103 | 168.666 |  |  |
|  | Total | 35219.257 |  |  |  |
| ECSB | Between Groups | 381.232 | 127.077 | 1.086 | 0.356 |
|  | Within Groups | 24114.083 | 117.059 |  |  |
|  | Total | 24495.314 |  |  |  |

Key: CBAB; Controlling bleeding after birth, HBB; Helping babies breath, ECSB; Essential Care for Small Babies

### Participants’ skills retention scores

Five skills areas were assessed including, three for the care of the mother i.e. the manual removal of placenta, active management of the third stage of labour, and administration of magnesium sulphate, and two for the care of the newborn i.e. helping babies breathe and feeding with a nasal gastric tube. Participants’ follow-up mean skills (SD) score was highest for manual removal of placenta {68.5 (16.1)} and lowest for feeding with nasal gastric tube {66.4 (14.6)}. The results showed that there was a drop in mean skills scores in all areas assessed with the highest drop observed in the administration of magnesium sulphate {-17.1 (18.7)} and lowest in helping babies breathe {-11.1 (11.0)}.

A paired *t*-test was performed to assess the differences between the follow-up mean skills scores and the post-test skills mean scores (**Table 6**) to determine whether the changes in skills scores were statistically significant. In all skills areas assessed, a statistically significant drop in scores was observed. Specifically, there was a significant drop in mean skill scores for manual removal of placenta (*t*=13.427, *p*<0.001), active management of third stage of labour (*t*=15.840, *p*<0.001), administration of magnesium sulphate (*t*=13.210, *p*<0.001), helping babies breathe (*t*=14.609, *p*<0.001), and feeding with nasal gastric tube (t=16.113, *p*<0.001)

**Table 6.** Mean scores differences between follow-up and post-test skills scores.

|  | Post-test |  | Follow-up |  | Difference |  | Paired *t*-test |  |  |
| --- | --- | --- | --- | --- | --- | --- | --- | --- | --- |
|  | Mean | SD | Mean | SD | Mean | SD | 95%CI | *t* | p value |
| MRP | 83.6000 | 7.88688 | 68.4905 | 16.07912 | -15.1095 | 16.30723 | 12.89112, 17.32793 | 13.427 | <0.001 |
| AMTSL | 83.9952 | 8.15255 | 66.9571 | 14.47694 | -17.0381 | 15.58795 | 14.91754, 19.15865 | 15.840 | <0.001 |
| AdMgSO<sub>4</sub> | 83.6268 | 9.49162 | 66.6048 | 20.56920 | -17.0766 | 18.68859 | 14.52805, 19.62506 | 13.210 | <0.001 |
| HBB | 78.6190 | 7.54747 | 67.5095 | 14.45178 | -11.1095 | 11.02031 | 9.61034, 12.60871 | 14.609 | <0.001 |
| FNGT | 78.8571 | 7.13027 | 66.4048 | 14.56886 | -12.4524 | 11.19895 | 10.92890, 13.97586 | 16.113 | <0.001 |

Key: MRP; Manual removal of placenta, AMTSL; active management of third stage of labour, AdMgSO<sub>4</sub>; Administration of magnesium sulphate, HBB; Helping babies breathe, FNGT; Feeding with nasogastric tube, CI; confidence interval

An independent *t*-test (**Table 7**) was also performed to compare the mean skills drop scores between participants working in urban and rural areas. Participants working in rural area had a statistically significant higher drop in mean skills scores for manual removal of placenta (F=14.917, *p*<0.001), and for active management of the third stage of labour (F=6.271, *p*=0.013).

**Table 7.** Univariate analysis of variance of skills according to residential area.

| Measure | Urban |  | Rural |  | Independent *t*-test |  |  |
| --- | --- | --- | --- | --- | --- | --- | --- |
|  | Mean | SD | Mean | SD | F (1, 208) | *p* value | η<sup>2</sup> |
| MRP | -11.6 | 15.3 | -20.2 | 16.5 | 14.917 | <0.001 | 0.067 |
| AMTSL | -14.8 | 16.1 | -20.2 | 14.3 | 6.271 | 0.013 | 0.029 |
| AdMgSO<sub>4</sub> | -15.4 | 17.5 | -19.4 | 20.2 | 2.356 | 0.126 | 0.011 |
| HBB | -12.1 | 11.6 | -9.7 | 10.0 | 2.367 | 0.125 | 0.011 |
| FNGT | -13.1 | 12.1 | -11.5 | 9.8 | 1.028 | 0.312 | 0.005 |

Key: MRP; Manual removal of placenta, AMTSL; active management of third stage of labour, AdMgSO<sub>4</sub>; Administration of magnesium sulphate, HBB; Helping babies breathe, FNGT; Feeding with nasogastric tube, CI; confidence interval η<sup>2</sup>; Eta Squared

**Table 8.**
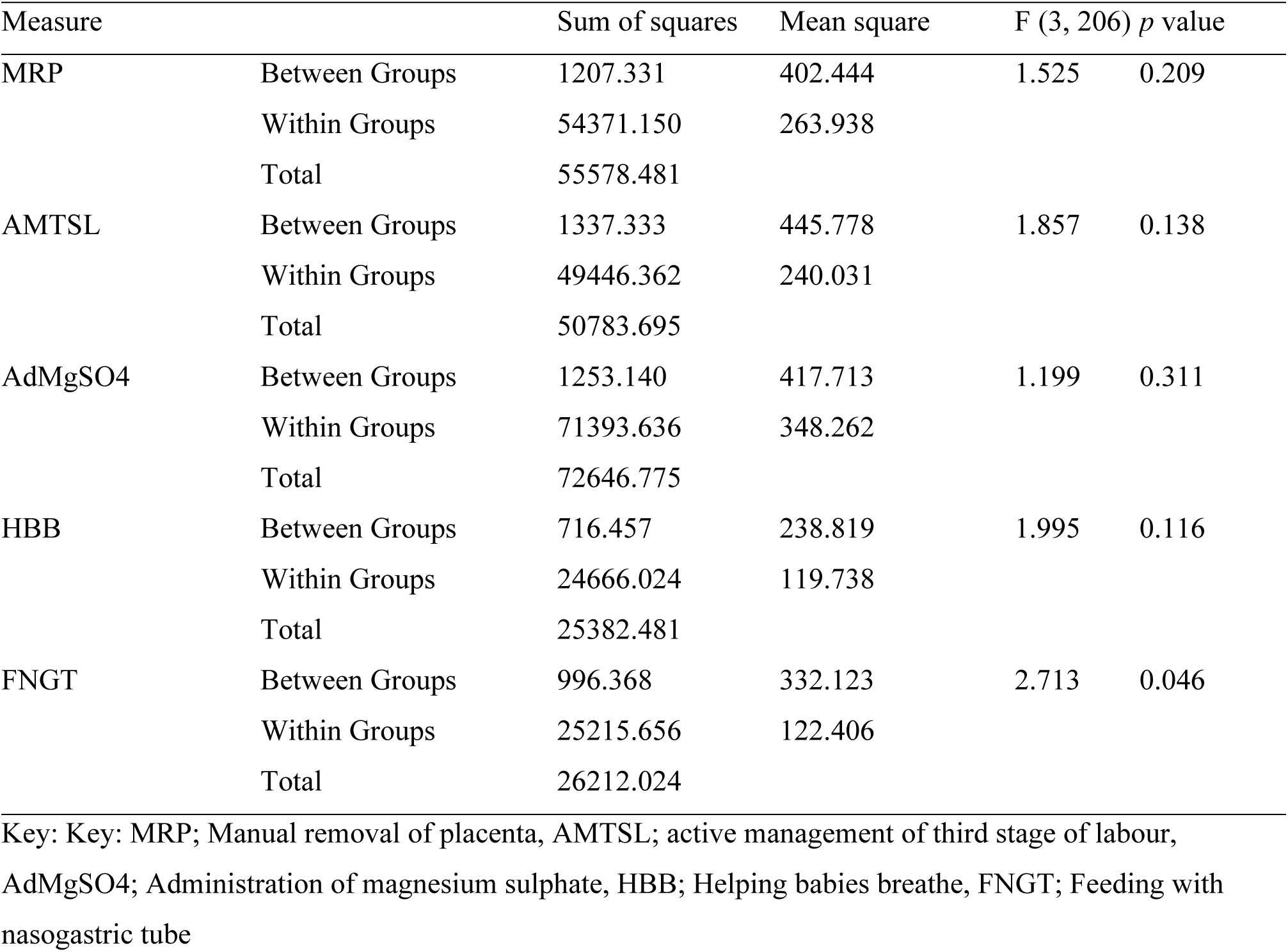
One Way Analysis of Variance of skills by level of professional education.

### Factors influencing the knowledge and skills retention

#### 1. Learning materials

##### a. Availability of learning materials

Participants reported the availability of learning materials such as handouts, and reference books as a crucial factor for knowledge and skills retention. Learning materials allow learners to review, reinforce, and apply the concepts and skills they have acquired in the classroom. Handouts and reference books can also serve as feedback, guidance, and self-assessment sources for learners. By having access to learning materials, learners can enhance their understanding, deepen their memory, and transfer their knowledge to new situations.

> *“For example, when you are given out a handout, and revise every time, it will help you to build experience. Although you may not be able to attend to a patient physically but it will assist to read the references and help put your knowledge intact.” (FGD Tanga)*

##### b. Topic revision in CMEs

Participants highlighted the importance of topic revision during the Clinical Medical Education sessions (CMEs) as one of the factors enhancing knowledge and skills retention. Topic revision is a factor for knowledge retention because it helps learners consolidate their learning and recall it more easily. Moreover, revising topics after a gap can strengthen memory traces and make them more resistant to forgetting.

> *“We normally have learning sessions at our facility; we have a teaching plan. Therefore, every Wednesday we prepare a lesson. We prepare maybe this week it will be eclampsia, next week it will be PPH, and we learn through those also we get experience.” (FGD Tanga)*

##### c. Establishment of a learning corner

Participants reported the establishment of a learning corner as another factor for knowledge retention because it provides a space where learners can actively engage with the content and review the material at their own pace. A learning corner can also help learners apply their knowledge to real-world scenarios and problems, which can increase their confidence and competence. Participants declared the presence of a learning corner that they use to refresh their knowledge and skills in the provision of maternal and newborn care.

> *“There are times, you may have what we call ‘learning corner’. Sometimes you may not have a client and the labour ward is calm no any challenge. Therefore, you go to the small room where there is a learning corner there we have manikins ‘Mama Natalie, Mama Besiye, Mama Yu’ and other manikins. Therefore, we are able to learn from those manikins at the end of the day you can come across a real case or real patient and you can recall what you have gained or this and we transfer (the knowledge and skills) to the real patient” (IDI, GRRH)*

#### 2. Mentorship and supportive supervision

Participants highlighted that they need to have mentorship and supervision by being followed up to ensure they master the content that has been taught during the training. Mentorship and supportive supervision were reported to help them learn from experienced mentors, receive constructive feedback, and improve their skills and performance. Mentorship and supportive supervision also foster a culture of learning and collaboration, where providers can share their knowledge and insights with each other.

> *“Mentorship should be done and supervision from our experts from CHMT and when they do supervision several times, they bring you back to class and do the required process which is needed in the trained issues” (IDI, Kasamwa)*

#### 3. Practice

##### a. Frequent practice of the procedure

Participants responded that having opportunities for frequent practice of the procedure tends to enhance knowledge and skills retention. Frequent practice of the procedure is a crucial factor for knowledge and skills retention. Frequent practice helps them to consolidate their memory, build confidence and refine their performance, and overcome challenges. Without frequent practice, learners may forget the steps of the procedure, lose confidence, and make errors.

> *“Okay, the big thing is that we should not get tired of practicing at all! When you do…you practice a lot it builds…builds confidence and ability in a sense that you will not forget because you have touched it with your own hands meaning it will remain in your mind what you did…therefore every day the weapon is to practice nothing else.” (FGD Muheza)*

##### b. Attending cases following the topics that they have been trained

Participants responded that attending cases following the topics that they have been trained in is one factor that can influence their knowledge retention and performance. By attending cases, they can reinforce their learning, gain confidence and feedback, and adapt to different contexts and scenarios.

> *“…it helps to remain with knowledge when we meet a case several times. For example, at that time we received training on manual removal of the placenta. Within that same week, I went and attended the case, and I found that I was able to manage it. Therefore, when we get several cases, we keep on learning.” (FGD Tanga)*

#### 4. Staff rotation/shifting

Participants have further reported staff rotation or shifting as another factor that positively and negatively influences knowledge and skills retention. Staff rotation or shifting is the practice of assigning healthcare providers to different tasks or positions within a facility. This can have various benefits and challenges for knowledge and skills retention. On the other hand, staff rotation can lead to some risks for knowledge and skills retention, such as loss of hands-on skills, reduced efficiency, and increased training costs (need for re-training).

> *“…when shifted to another unit loses the skills, because if you do not practice for a long time…as time goes on you keep on forgetting (IDI, Katavi)”*

> *“First it is supposed that midwife work in the same maternity unit, and after taking training may be of helping the baby to breathe and then you take to the surgical ward or medical ward, there you have not helped, therefore after training the first thing is to remain in the maternity unit” (IDI, Ilembo)*

> *“Skills, to retain skills is when someone is working in the same unit meaning for example after training, then works for at least for some time maybe six months. He/she keeps working in the labour unit where training was conducted helps to gain the skills and retain them for a longer period. But if after training, she/he is shifted from labour and goes to maybe OPD…when she/he goes back the skills will be gone since has not practised them…” (IDI, Kasamwa)*

#### 5. Training

Training is another factor for knowledge and skills retention that the participants have described. Training can take various forms, such as retaining on-the-job training, teaching others, self-directed learning through individual revision, receiving updates on evidence based practice etc. Training through these various forms can enhance knowledge and skills retention by increasing the confidence, competence, and commitment of healthcare providers.

> *“…doing sessions every week and we do on-job training…For example, when we receive an emergency PPH case, when you provide the care, you are with fellow providers…and we teach each other” (IDI, Buga)*

> *“In my opinion, I think this training should be conducted several times. Therefore, if this group receives training after some time you prepare training for another group. It will help people at work very much. You know when you receive on-the-job training again it helps a lot and builds morale and competence.” (FGD, Tanga).*

> *“…because, for you to be able to have more education and stick in your head it is when you teach others. When you keep on teaching others things and things keep on sticking into your head therefore eventually your competence will be improved and people will receive high-quality services.”(FGD, Muheza)*

#### 6. Availability of equipment

Participants also highlighted the availability of equipment as a factor influencing knowledge and skills retention among healthcare providers. This is because it enables providers to apply their learning in practice and improve their performance. However, the availability of equipment alone is not sufficient for knowledge and skills retention. Receiving adequate training needs to go along with the availability of equipment.

> *“…for example, if you have enough equipment, they also assist you to give good services with the required skills. But if you have scarce equipment also it becomes a challenge to give quality services and remain with the skills…but when you have enough equipment you provide good service properly.” (IDI, Buga).*

## Discussion

This was a mixed methods study aimed to assess factors influencing healthcare providers’ knowledge and skills retention following Helping Mothers Survive and Helping Babies Survive Training. Regarding knowledge, there was a significant drop in the four domains assessed including how to control bleeding after childbirth, managing pre-eclampsia, helping babies to breathe, and essential care for small babies. Similarly, there was a significant drop in the five skills areas assessed including the manual removal of placenta, active management of the third stage of labor, administration of magnesium sulphate, helping babies breathe, and feeding with a nasal gastric tube. A significant drop in specific knowledge and skills domains was higher among healthcare providers with lower professional education and those working in rural areas. Qualitatively, reported facilitators of knowledge and skills retention encompassed the availability of learning materials, frequent practice, mentorship and supportive supervision, on-job training, and staff rotation/shifting.

The current study demonstrates a statistically significant drop in knowledge and skills scores among healthcare providers following receiving the training on Helping Mothers Survive and Helping Babies Survive, indicating a decrease in knowledge and skills retention over time. A similar situation was observed in Uganda and Sudan where knowledge and skills regarding helping babies breathe among midwives fell by 12 months post-training [39, 40]. On the contrary, a previous study in the country which focused on one similar domain (Helping Mothers Survive Bleeding After Birth) reported that knowledge and simulated basic delivery skills decayed after nine months, while confidence and simulated obstetric emergency skills were largely retained [35]. Moreover, a study in Ethiopia reported an overall strong retention of knowledge and skills among health workers following maternal and newborn health training [41].

Various factors have been implicated in the maternal and neonatal care knowledge and skills retention including refresher training [40]. This is supported by a multi-country longitudinal study which showed that, after training, healthcare providers retain knowledge and skills for up to 12 months, the effect which likely enhanced by short repeat skills-training sessions, or, ‘fire drills’ [36]. Our findings from the qualitative arm emphasized the role of on-job training in enhancing essential maternal and newborn care knowledge and skills retention among healthcare providers.

Participants working in urban areas had a statistically significant higher drop in mean knowledge for managing pre-eclampsia and a statistically significant lower drop in mean knowledge for helping babies breathe. On the other hand, participants working in rural area had a statistically significant higher drop in mean skills for manual removal of placenta and active management of third stage of labor. This indicates differences experienced in knowledge and skills retention in the two settings. As reported in a previous study [42], there is scarcity of literature on professional education, training or continuous professional development in maternity care in remote and rural settings that can be compared with urban settings. However, the observed differences could be multifaceted and related to several factors, including differences in clinical exposure and experience, resource availability, and institutional practices [43, 44].

With regard to the level of professional education, certificate holder had a statistically significant higher drop in mean knowledge for pre-eclampsia compared to holders of diploma and degree. The differences in drop of mean skills was not significant. This implies that knowledge retention drop was much more experienced by the holders of certificate. This is supported by a previous study in Tanzania which found that professional qualification and experience in a maternity care settings are significant factors influencing not only nurses’ knowledge, but also skills [45]. The possible explanation is that healthcare workers with lower levels of professional education may have a weaker foundation in medical knowledge and clinical skills, leading to a more rapid decline in retained information [46].

Our study found that working in a hospital had a significantly higher drop in pre-eclampsia mean knowledge compared to those working in a dispensary. However, the situation was vice versa for skills for manual removal of placenta where healthcare providers working in dispensaries had a higher drop compared to those working in hospitals. This implies healthcare providers in hospitals and dispensaries had different experiences in various aspects of knowledge and skills retention. A previous multi-country study indicated that health care facility level was not a determinant of knowledge and skills retention among healthcare providers [36]. Various factors affecting each level of health facilities differently such as work experience, training type, training frequency, availability of equipment and facilities, and healthcare providers’ educational background may explain the observed differences in knowledge and skills retention [46]. Therefore, this highlights that health facility-specific factors should be considered to enhance healthcare providers’ knowledge and skills retention following training.

Our findings from the qualitative arm has highlighted the availability of learning materials, frequent practice, mentorship and supportive supervision, on-job training, and staff rotation/shifting as fractionators of knowledge and skills retention among healthcare providers. The identification of these factors emphasize that these are important factors to be considered to enhance knowledge and skills retention among healthcare providers. A discussed above, various studies support these findings [40, 43–46]. This implies that factors influencing knowledge and skills retention are multifaceted in nature and underscores the role of contextual considerations in addressing the phenomenon.

There are some strengths and limitations that are noteworthy in this study. Using a mixed design, the study draws its strength from the triangulation of information from both quantitative and qualitative arms. Findings from the qualitative arm were key in complementing the quantitative results. Moreover, the mix of urban and rural settings and various health facility levels provide a good sample representation of the study population. On the other hand, this study might have been influenced by social disability limitation, particularly with the qualitative arm as healthcare providers might want to provide desirable responses regarding factors influencing the knowledge and skills retention. However, they were assured of confidentiality and encouraged to provide honest responses to mitigate social desirability. However, we believe that this study provides invaluable insights regarding factors associated with knowledge and skills retention and among healthcare providers following the Helping Mothers Survive and Helping Babies Survive Training.

## Conclusion

This study highlights low level of knowledge and skills retention among healthcare providers following the Helping Mothers Survive and Helping Babies Survive training, with organizational factors being mainly associated. This underscores the need to address organizational barriers such as making available the learning materials, providing mentorship and supportive supervision, and conducting frequent on-job training to enhance knowledge and skills retention regarding maternal and child care among healthcare providers.

## Data Availability

Data is uploaded as supplementary information.

## Acknowledgements

We would like to thank all the HCPs who agreed to take part in the study and their willingness to share their insights regarding factors influencing competence retention. We extend our sincere gratitude to Laerdal Foundation for funding this study. Finally, we thank the assistant researchers who worked diligently in data collection. All regional and district authorities which gave us permission to conduct this study are acknowledged.

## Author Contributions

BEM, MR, FM and LM conceptualized the study, and were involved in analysis; BEM, MR, LM, and JA performed formal data analysis; BEM, JA, and RG were involved in writing the original draft and BEM, LM, MR, FM, RG, NK, JA contributed in writing – review and editing. Both authors revised and approved the final manuscript.

## Supporting Information

DATA SET ZIPPED FOLDER

